# Home and wild food procurement was associated with greater intake of fruits and vegetables during the COVID-19 pandemic in northern New England

**DOI:** 10.1101/2024.05.02.24306758

**Authors:** Ashley C. McCarthy, Ashleigh Angle, Sam Bliss, Farryl Bertmann, Emily H. Belarmino, Kelsey Rose, Meredith T. Niles

**Affiliations:** Department of Nutrition and Food Sciences, University of Vermont, Burlington, VT 05405; Gund Institute for Environment, University of Vermont, Burlington, VT 05405; Food Systems Program, University of Vermont, Burlington, VT 05405

**Keywords:** self-provisioning, gardening, food preservation, fishing, foraging, hunting, backyard livestock, fruit and vegetable intake, diet quality, meat intake, food insecurity, COVID-19

## Abstract

**Objective:** This study examined the effect of home and wild food procurement (HWFP) activities (i.e., gardening, hunting, fishing, foraging, preserving food, raising livestock, and raising poultry for eggs) on food security status, fruit and vegetable intake, and meat consumption.

**Design:** We used data collected in 2021 and 2022 through two statewide representative surveys (n = 2,001). Dietary intake was assessed using the Dietary Screener Questionnaire. We analyzed the data using linear regression, logistic regression, and ordinal logistic regression models.

**Setting:** Maine and Vermont, United States

**Participants:** 2,001 adults (18 years and older)

**Results:** Sixty-one percent of respondents engaged in HWFP activities; the majority of those gardened. Households engaging in most individual HWFP activities had greater odds of being food insecure. HWFP engagement was positively associated with fruit and vegetable consumption. Specifically, gardening was associated with an additional one cup-equivalent in fruit and vegetable consumption per week compared to respondents that did not garden. Furthermore, when exploring these relationships disaggregated by food security status, we find that this effect is stronger for food insecure households than food secure households. Respondents from households that hunted were more likely to eat wild game meat and also consumed red and white meat more frequently compared to households that did not hunt.

**Conclusion:** Overall, our results indicate potential public health and food security benefits from engaging in HWFP activities. Future research should continue to examine a full suite of HWFP activities and their relationship to diet, health, and food security.

## Introduction

Engagement in home and wild food procurement (HWFP) skyrocketed during the COVID-19 pandemic. For example, in the early days of the pandemic, authorities in 41 of 47 surveyed U.S. states reported increased license sales for spring turkey hunting ^(1)^. Participation in recreational fishing swelled too, in part because of the perceived safety of this outdoor activity; some anglers started to call it “social *fish-*tancing” ^(2–4)^. Vegetable seeds and mason jar lids became hard to find as new and existing gardeners bought them in preparation for augmenting their production and preservation capacities ^(5,6)^. Research shows people upped their participation in all these endeavors in part because they had more free time and wanted relief from the stress of the pandemic, but also because they were worried about having enough food ^(7,8)^.

Growing and wild-harvesting one’s own food can serve as coping mechanisms for dealing with food insecurity ^(9,10)^. A 2020 survey of Vermonters found that food-insecure households were more likely to engage in a suite of HWFP activities except gardening in the first six months of the pandemic as compared to food-secure households ^(11)^. Furthermore, there is some evidence that these practices work to improve food security: among households that experienced food insecurity in the early months of the COVID-19 pandemic, those that engaged in HWFP were significantly more likely to become food secure a year into the pandemic ^(12)^.

Evidence of HWFP’s ability to improve diet and nutrition during the pandemic is less established. Across high-income societies, many studies have found an association between gardening and greater fruit and vegetable intake ^(13)^, though this effect has been identified in some cases as only occurring for food secure households ^(11)^. Yet many of these studies do not have non-gardening control groups ^(14–17)^, or they are correlational, precluding examination of causal relationships ^(18,19)^ - it may not be that households eat more vegetables because they garden, but the other way around: that they garden because they like eating vegetables. A recent randomized control trial found that the group that received a gardening intervention increased their vegetable consumption compared to the control group, by 0.67 servings per day ^(20)^. In interviews, gardeners said they ate more vegetables because of increased availability, better taste, trying new dishes, pride in their homegrown food, not wanting to waste it, and emotional connections with gardens and their plants ^(20)^.

Across high income countries, research on the dietary impacts of HWFP activities is limited to studies on gardening and associations between fishing and omega-3 fatty acid levels ^(21,22)^. Rigorous research relating hunting, foraging, fishing, and raising livestock to the intake of relevant foods and nutrients comes entirely from the Global South or remote Indigenous communities ^(23–27)^. The few studies that examine a full suite of HWFP activities as a unified category ^(28–30)^ do not address diet, nutrition, or food security.

This study fills these gaps by exploring a suite of HWFP activities in a high-income country and looks at the relationships between HWFP activities and food security and dietary intake. We use survey data from a representative sample (n=2,001) of residents in Maine and Vermont, the two U.S. states with the greatest share of residents living in rural areas, to assess how household-level HWFP engagement relates to food security status and intake of foods that certain HWFP activities produce: fruits, vegetables, and game meat. Thus, our analysis goes beyond gardening to get a population-level view of the dietary implications of most HWFP activities. Unlike available studies on gardening and vegetable intake, we explore the differential effect that engaging in HWFP may have for food-secure versus -insecure households. Previous research has demonstrated that food security status can influence the potential benefits accrued through engagement in these activities ^(11,31)^.

We evaluate the following hypotheses:

- H1: Engagement in HWFP is positively associated with food insecurity.
- H2a: Engagement in gardening, foraging, and preserving food are associated with higher fruit and vegetable intake.
- H2b: Food insecure households that gardened, foraged, and preserved food will have lower fruit and vegetable consumption, as compared to food secure households that engaged in these activities.
- H3a: Engagement in hunting is associated with eating game meat more frequently.
- H3b: Engagement in hunting is associated with eating red meat and white meat less frequently.
- H3c: Food insecure households that hunted will have greater odds of eating game meat, as compared to food insecure households that did not hunt.

By gaining a fuller understanding of HWFP’s relationship with food security and diet outcomes, we hope to identify the potential of HWFP as a strategy to improve food security and diet quality.

## Methods

### Survey Development and Sampling Strategy

Data were collected through two surveys in the US states of Maine and Vermont. The first survey was conducted from March to June 2021 (n=988) and the second survey was conducted from April to May 2022 (n=1,013). The survey instrument was initially developed in March 2020 by the National Food Access and COVID research Team (NFACT) ^(32)^ and then expanded for 2021 and 2022 to include more questions about HWFP activities ^(33)^. The survey included sections on food sourcing, HWFP, food security, diet intake, health outcomes and behaviors, experiences during the COVID-19 pandemic, and demographic information. Institutional Review Board approval was obtained prior to data collection. Participants were recruited through Qualtrics (Provo, UT) research panels and completed an online survey. We used sample quotas to ensure our respondents were representative of the racial and ethnic distributions of the states based on the population profiles from the American Community Survey ^(34)^. Respondents were anonymous in the data collection process.

### Variables for Analysis

We used four categories of variables in our analyses: food security status, engagement in HWFP activities, dietary intake, and demographic information (Table 1).

**Table 1.** Complete list of variables included in the analysis.

| **Variable Name** | **Question(s)** | **Scale** |
| --- | --- | --- |
| Food Security Status | USDA 6-item food security model for past 12 months | 0 = Food Secure; 1 = Food Insecure |
| **Dietary Quality Variables** |  |  |
| Fruit and Vegetable Intake | Predicted intake of fruits and vegetables including legumes and excluding French fries based on DSQ | Cup Equivalents Per Day |
| Fruit Intake | Predicted intake of fruits (including 100% pure fruit juice) based on DSQ | Cup Equivalents Per Day |
| Vegetable Intake | Predicted intake of vegetables excluding French fries based on DSQ | Cup Equivalents Per Day |
| Game Meat Consumption | During the past month, how often did you eat wild game meat such as venison, wild turkey, pheasant, or bear? | 0 = Did Not Consume; 1 = Did Consume |
| Red Meat Consumption | During the past month, how often did you eat red meat, such as beef, pork, ham, or sausage? | 1 = Never; 2 = 1 time in the last month; 3 = 2-3 times in the last month; 4 = 1 time per week; 5 = 2 times per week; 6 = 3-4 times per week; 7 = 5-6 times per week; 8 = 1 time per day; 9 = 2 or more times per day |
| White Meat Consumption | During the past month, how often did you eat white meat, such as chicken and turkey? |  |
| **Home and Wild Food Procurement (HWFP) Variables** |  |  |
| Gardening<br>Foraging<br>Fishing<br>Hunting<br>Raising Livestock<br>Raising Poultry for Eggs<br>Preserving Food | Has your household engaged in any of these activities in the following in the last 12 months? | 0 = No; 1 = Yes |
| Any HWFP | Variable created based on respondent indicating that they engaged in any of the individual HWFP activities | 0 = No; 1 = Yes |

| **Demographic Variables** |  |  |
| --- | --- | --- |
| Annual Household Income | Which of the following best describes your household income range before taxes? [2021 survey asked about 2019 income; 2022 survey asked about 2021 income] | 0 = Household income < \$50,000<br>1 = Household income ≥ \$50,000 |
| College Degree | What is the highest level of formal education that you have completed? | 0 = Less than associate's degree<br>1 = Associate's or higher |
| Age | In what year were you born? | 0 = Under 65; 1 = 65 and older |
| Race | What is your race? Check all that apply. | Response options include:<br>American Indian/Alaska Native;<br>Asian Indian; Black or African American; Chamorro; Chinese; Filipino; Japanese; Korean; Native Hawaiian; Samoan; Vietnamese; White |
| Ethnicity | Are you of Hispanic, Latino, or Spanish origin? | 1 = No, not Hispanic, Latino, or Spanish origin; 2 = Yes, Mexican, Mexican American, Chicano; 3 = Yes, Puerto Rican; 4 = Yes, Cuban; 5 = Yes, Hispanic, Latino, or Spanish origin |
| Race/Ethnicity Binary | Based on responses to race and ethnicity questions above | 0 = BIPOC; 1 = non-Hispanic White |
| Gender Identity | Which of the following best describes your gender identity? | 0 = Male; 1 = Female; 3 = Another Gender Identity |
| Rural/urban classification | Rural or urban classification based on zip code responses and RUCA codes | 0 = Rural; 1 = Urban |
| Job Disruption | Have you or anyone in your household experienced a loss of income, reduction of hours, furlough or job loss since the COVID-19 outbreak began March 11th, 2020? | 0 = No; 1 = Yes |

Food security status was measured using the US Department of Agriculture 6-item short-form food security module ^(35)^. Respondents were asked to assess whether their households had sufficient economic access to food during the 12 months prior to taking the survey. Following the standard protocol for calculating food insecurity, respondents who responded affirmatively to two or more questions were classified as food insecure.

Engagement in HWFP activities was assessed through a series of questions about whether anyone in the respondent’s household had gardened, hunted, fished, foraged, raised livestock for meat or milk, raised poultry for eggs, or preserved food during the 12 months prior to taking the survey. We analyzed engagement in any HWFP activity and engagement in specific activities.

Dietary intake was measured using the Dietary Screener Questionnaire (DSQ), a standardized and validated instrument to identify the frequency of intake of selected foods during the previous 30 days ^(36)^. For each of the 26 items included in the screener, respondents chose one of nine frequency options ranging from never to multiple times per day. We added a question to the screener to measure frequency of game meat (e.g., venison, wild turkey or duck, pheasant, bear, etc.) consumption, since game meat is not included in the DSQ. Although the DSQ does not measure quantity consumed, scoring algorithms provided by the National Cancer Institute (NCI) were used to convert screener responses to estimate daily intake (in cup equivalents or cup-eq) for fruits and vegetables ^(37)^. Red meat, white meat, and game meat are reported on a consumption frequency basis and as a binary variable indicating absence or presence of any consumption.

Individual demographics include age, gender identity, race, ethnicity, and education level. Household demographics included annual household income, job disruption during the COVID-19 pandemic, and zip code. We used respondent zip codes and the Rural-Urban Commuting Area (RUCA) code system to classify respondent geography as urban or rural ^(38,39)^.

### Statistical Analysis

To examine the relationship between HWFP engagement and food security status, we used logistic regression models, reporting the odds ratios. We constructed separate models for each individual HWFP activity, as well as a combined model that included all activities to understand interactions between them.

To examine the relationship between HWFP engagement and fruit and vegetable intake, we used linear regression models, reporting the coefficients. In these analyses, we looked at the three HWFP activities that produce fruits and vegetables: gardening, foraging, and preserving food.

Once again, we used separate models that looked at each individual activity’s relationship to fruit and vegetable consumption, plus a combined model that included all three activities to understand any interactions between them. We also examined the relationship by food security status by running separate analyses only including food-secure households and only including food-insecure households.

To examine the relationship between hunting and frequency of meat consumption, we used logistic regression models for game meat and ordinal logistic models for red meat and white meat, reporting odds ratios. Due to the distribution of the data, we had to analyze game meat consumption as a binary variable (any consumption vs no consumption) rather than looking at a more detailed range in consumption frequency. We also examined the relationship by food security status by running separate analyses only including food-secure households and only including food-insecure households.

All analyses controlled for household income, age, education level, race/ethnicity, rurality, gender identity, and job disruptions during the pandemic. We also included a dummy variable in all analyses to control for survey year (2021 or 2022). All analyses used survey weights to correct for income distribution because our sample over-represents low-income households. We report statistical significance as p ≤ 0.05. All analyses were done in Stata v17.0 ^(40)^.

## Results

Table 2 presents sample characteristics. There were no major differences in outcomes by state; thus, Maine and Vermont respondents were combined for this analysis. The sample reflects the adult populations of Maine and Vermont with respect to race and ethnicity but over-represents low-income households (which we control for in models using weighting).

**Table 2.** Demographic characteristics of survey respondents (n=2,001)

| Category | Characteristic | Sample |  | Population<sup>1</sup> |
| --- | --- | --- | --- | --- |
|  |  | n | % | % |
| Age | < 65 | 1,576 | 78.8 | 76.7 |
|  | ≥ 65 | 425 | 21.2 | 23.3 |
| Gender | Male | 657 | 32.8 | 49.4 |
|  | Female | 1,308 | 65.4 | 50.6 |
|  | Other Gender | 28 | 1.4 | n/a |
| Race/Ethnicity | Non-Hispanic White | 1,827 | 91.3 | 93.1 |
|  | BIPOC | 157 | 7.9 | 6.9 |
| Education Level | No College Degree | 1,052 | 52.6 | 54.4 |
|  | College Degree | 949 | 47.4 | 45.6 |
| Household Income | < \$50,000/year | 973 | 48.6 | 38.8 |
| | ≥ \$50,000/year | 1,027 | 51.3 | 61.2 |
| Rurality | Urban | 853 | 42.7 | 46.5 |
|  | Rural | 1,143 | 57.1 | 53.5 |
| Job Disruption | Yes | 1,230 | 61.5 | n/a |
|  | No | 770 | 38.5 | n/a |

<sup>1</sup>Based on US Census Bureau 2017–2021 American Community Survey estimates and 2010 Decennial Census data for Maine and Vermont Categories do not sum to 100% due to missing data

Among all respondents, 61.0% indicated that their household had engaged in an HWFP activity within 12 months of completing the survey. The most common activity was gardening (45.9%), followed by preserving food (29.6%), and fishing (15.9%) (Figure 1). The least common activities were raising livestock for meat or milk and raising poultry for eggs. Among respondents who engaged in HWFP, 55.5% reported engaging in two or more HWFP activities. For example, most (79.4%) foraging households also gardened and 61.4% of hunting households also fished.

**Figure 1.**
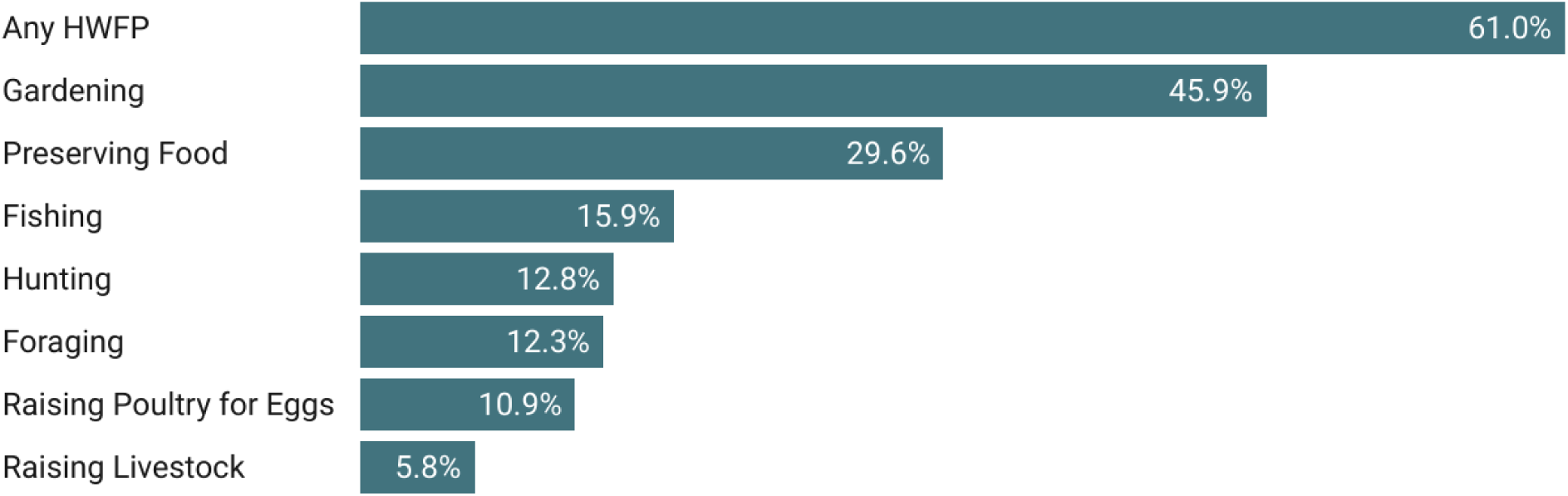
Share of respondents that indicated they engaged in HWFP activities in the last 12 months.

### HWFP Engagement and Food Security Status

Among the respondents who completed the food security module (n = 1,890), 34.9% were food insecure at some point in the 12 months prior to taking the survey while 65.1% were food secure.

We used separate logistic regression models to examine the relationships between food security status and engaging each individual HWFP activity or engaging in any HWFP (H1). We found that households that engaged in foraging (OR=1.61, p=0.006), hunting (OR=1.64, p=0.003), fishing (OR=1.60, p=0.005), raising livestock (OR=2.71, p<0.001), and raising poultry for eggs (OR=2.07, p<0.001) were more likely to be food insecure than households that did not do these activities (Table 3; full results of each model in supplementary material tables S1-S8). Combining all HWFP activities together in a single model, we find no statistically significant relationship between HWFP engagement and food security status except for those that raised poultry for eggs (OR=1.56; p=0.041), who were significantly more likely to be food insecure (Table 4).

**Table 3.** Summary of results from separate logistic regression models predicting the odds of food insecurity by overall and specific HWFP activity engagement.

| Activity | Odds Ratio | Standard Error | p-value | 95% CI |  |
| --- | --- | --- | --- | --- | --- |
| Any HWFP | 1.19 | 0.141 | 0.132 | 0.948 | 1.504 |
| Gardening | 1.06 | 0.123 | 0.606 | 0.846 | 1.331 |
| Preserving Food | 1.22 | 0.159 | 0.128 | 0.945 | 1.574 |
| **Foraging** | **1.61** | **0.275** | **0.006** | **1.149** | **2.248** |
| **Hunting** | **1.64** | **0.277** | **0.003** | **1.177** | **2.283** |
| **Fishing** | **1.60** | **0.264** | **0.005** | **1.156** | **2.208** |
| **Raising Livestock** | **2.71** | **0.733** | **0.000** | **1.592** | **4.602** |
| **Raising Poultry for Eggs** | **2.07** | **0.390** | **0.000** | **1.433** | **2.997** |

Odds ratios higher than 1.00 indicate a greater odds of food insecurity. Each model also included control variables for age, gender identity, race/ethnicity, education level, income, rurality, job disruption, and survey year. Statistically significant ( $p < 0.05$ ) results are bolded for emphasis.

**Table 4.** Results of logistic regression model predicting the combined effects of HWFP engagement on food security status.

| Variable | Odds Ratio | Standard Error | p-value | 95% CI |  |
| --- | --- | --- | --- | --- | --- |
| Gardening | 0.85 | 0.110 | 0.204 | 0.656 | 1.094 |
| Preserving Food | 0.99 | 0.148 | 0.987 | 0.745 | 1.335 |
| Foraging | 1.27 | 0.239 | 0.197 | 0.882 | 1.839 |
| Hunting | 1.08 | 0.217 | 0.716 | 0.725 | 1.599 |
| Fishing | 1.24 | 0.242 | 0.272 | 0.845 | 1.818 |
| Raising Livestock | 1.71 | 0.535 | 0.089 | 0.922 | 3.154 |
| **Raising Poultry for Eggs** | **1.56** | **0.339** | **0.041** | **1.018** | **2.390** |
| Race/Ethnicity | 0.75 | 0.154 | 0.156 | 0.497 | 1.118 |
| Gender Identity | 1.08 | 0.136 | 0.536 | 0.845 | 1.383 |
| **Age** | **0.32** | **0.053** | **0.000** | **0.228** | **0.438** |
| **Education Level** | **0.52** | **0.063** | **0.000** | **0.408** | **0.656** |
| **Household Income** | **0.25** | **0.031** | **0.000** | **0.199** | **0.322** |
| Rurality | 1.05 | 0.121 | 0.670 | 0.839 | 1.315 |
| **Job Disruption** | **3.54** | **0.417** | **0.000** | **2.814** | **4.462** |
| **Survey Year** | **1.64** | **0.197** | **0.000** | **1.297** | **2.076** |

Odds ratios higher than 1.00 indicate a greater odds of food insecurity. Statistically significant ( $p < 0.05$ ) results are bolded for emphasis.

### HWFP Engagement and Fruit and Vegetable Intake

Respondents reported an average fruit intake of 0.85 cup-eq/day and an average vegetable intake of 1.47 cup-eq/day, for a combined fruit and vegetable intake of 2.32 cup-eq/day. The 2020-2025 Dietary Guidelines for Americans (DGA) recommend that most adults consume 2 cup-eq of fruit per day and 2.5 to 3.5 cup-eq of vegetables per day ^(41)^. Fewer than 2% of the respondents in our sample met these DGA recommendations for either fruit intake or vegetable intake.

When looking at fruit and vegetable intake in relation to engagement in relevant HWFP activities (H2a), we found that gardening, foraging, and preserving food were significantly positively associated with fruit, vegetable, and combined fruit and vegetable intake in all instances except fruit intake and foraging, for which no relationship was identified (Table 5; full results of each model in supplementary material tables S9-S17). Specifically, respondents who gardened had a 0.09 cup-eq/day greater intake for vegetables (p<0.001), 0.07 cup-eq/day for fruits (p<0.001), and 0.15 cup-eq/day for combined fruit and vegetable intake (p<0.001). These daily amounts equate to an additional 1.1 cup-eq of combined fruit and vegetable intake per week. Similarly, preserving food was associated with a 0.05 cup-eq/day greater intake of both vegetables and fruits (p=0.028). Foraging was significantly associated with a 0.08 cup-eq/day greater intake for vegetables (p=0.012) and a 0.11 cup-eq/day greater intake for fruits and vegetables combined (p=0.038), more than an additional ¾ cup-eq over the course of a week. Exploring the relationship of these activities together in a single model demonstrated that collectively only gardening is associated with greater fruit and vegetable intake (0.14 cup-eq/day increase for fruits and vegetables combined (p<0.001)) (Table 6; full results of each model in supplementary material tables S18-S20).

**Table 5.** Summary of results from separate linear regression models predicting the effects of HWFP engagement on daily fruit and vegetable intake (cup equivalents).

| **HWFP Activity** | **Diet Factor** | **Coefficient** | **Standard Error** | **p-value** | **95% CI** |  |
| --- | --- | --- | --- | --- | --- | --- |
| Gardening | **Vegetable Intake** | **0.09** | **0.020** | **0.000** | **0.049** | **0.130** |
|  | **Fruit Intake** | **0.07** | **0.019** | **0.000** | **0.031** | **0.104** |
|  | **Fruit and Vegetable Intake** | **0.15** | **0.033** | **0.000** | **0.087** | **0.217** |
| Preserving Food | **Vegetable Intake** | **0.05** | **0.022** | **0.028** | **0.005** | **0.093** |
|  | **Fruit Intake** | **0.05** | **0.021** | **0.018** | **0.009** | **0.093** |
|  | **Fruit and Vegetable Intake** | **0.09** | **0.037** | **0.019** | **0.014** | **0.160** |
| Foraging | **Vegetable Intake** | **0.08** | **0.030** | **0.012** | **0.017** | **0.138** |
|  | Fruit Intake | 0.03 | 0.028 | 0.248 | -0.023 | 0.089 |
|  | **Fruit and Vegetable Intake** | **0.11** | **0.051** | **0.038** | **0.005** | **0.210** |

Each model also included control variables for age, gender identity, race/ethnicity, education level, income, rurality, job disruption, and survey year. Statistically significant ( $p < 0.05$ ) results are bolded for emphasis.

**Table 6.** Summary of results for linear regression models predicting the combined effects of HWFP engagement on daily fruit and vegetable intake (cup equivalents).

| **Diet Factor** | **HWFP Activity** | **Coefficient** | **Standard Error** | **p-value** | **95% CI** |  |
| --- | --- | --- | --- | --- | --- | --- |
| Vegetable Intake | **Gardening** | **0.08** | **0.022** | **0.000** | **0.034** | **0.120** |
|  | Foraging | 0.04 | 0.034 | 0.190 | -0.022 | 0.059 |
|  | Preserving Food | 0.01 | 0.024 | 0.627 | -0.035 | 0.059 |
| Fruit Intake | **Gardening** | **0.06** | **0.021** | **0.005** | **0.018** | **0.099** |
|  | Foraging | 0.00 | 0.030 | 0.981 | -0.059 | 0.060 |
|  | Preserving Food | 0.03 | 0.023 | 0.212 | -0.016 | 0.074 |
| Fruit and Vegetable Intake | **Gardening** | **0.14** | **0.037** | **0.000** | **0.064** | **0.207** |
|  | Foraging | 0.04 | 0.056 | 0.421 | -0.064 | 0.154 |
|  | Preserving Food | 0.03 | 0.040 | 0.487 | -0.050 | 0.207 |

Each model also included control variables for age, gender identity, race/ethnicity, education level, income, rurality, job disruption, and survey year. Statistically significant ( $p < 0.05$ ) results are bolded for emphasis.

We also explored the relationship between HWFP engagement and combined fruit and vegetable intake by food security status (H2b) (Table 7; full results of each model in supplementary material tables S21-S24). Engaging in any HWFP activity is associated with greater combined fruit and vegetable intake among food insecure households (b=0.14; p=0.005), but not food secure households (p=0.074). Participating in gardening is associated with greater combined fruit and vegetable intake among both food secure (b= 0.12; p=0.006) and food insecure households (b= 0.22; p < 0.000). For food insecure households, this is the equivalent of nearly 1/4 cup greater fruit and vegetable intake per day, or more than a cup and a half a week. Foraging (b= 0.18; p=0.009) and food preservation (b=0.10, p=0.050) are also both associated with greater combined fruit and vegetable intake, but only for food secure households. Looking at the combined effect of all three relevant HWFP activities in a single model, only gardening among food insecure households had a statistically significant effect on combined fruit and vegetable intake (Table 8). Among food insecure households, gardening was associated with a 0.23 cup-eq/day greater fruit and vegetable intake (p<0.000), more than a cup and a half a week.

**Table 7.** Summary of results from separate linear regression models predicting the effects of HWFP engagement on daily combined fruit and vegetable intake (cup equivalents), by food security status.

| HWFP Activity | Food Security Status | Coefficient | Standard Error | p-value | 95% CI |  |
| --- | --- | --- | --- | --- | --- | --- |
| Any HWFP | Food Secure | 0.08 | 0.043 | 0.074 | -0.017 | 0.150 |
|  | **Food Insecure** | **0.14** | **0.050** | **0.005** | **0.113** | **0.327** |
| Gardening | **Food Secure** | **0.12** | **0.043** | **0.006** | **0.035** | **0.200** |
|  | **Food Insecure** | **0.22** | **0.054** | **0.000** | **0.100** | **0.327** |
| Preservation | **Food Secure** | **0.10** | **0.048** | **0.050** | **0.000** | **0.192** |
|  | Food Insecure | 0.07 | 0.060 | 0.245 | -0.048 | 0.188 |
| Foraging | **Food Secure** | **0.18** | **0.069** | **0.009** | **0.045** | **0.320** |
|  | Food Insecure | 0.02 | 0.079 | 0.851 | -0.142 | 0.172 |

Each model also included control variables for age, gender identity, race/ethnicity, education level, income, rurality, job disruption, and survey year. Statistically significant ( $p<0.05$ ) results are bolded for emphasis.

**Table 8.** Linear regression results predicting the combined effects of gardening, food preservation, and foraging engagement on daily fruit and vegetable intake (cup equivalents), by food security status.

| Fruit and Vegetable Intake | Coefficient | Standard Error | p-value | 95% CI |  |
| --- | --- | --- | --- | --- | --- |
| Food Secure |  |  |  |  |  |
| Gardening | 0.087 | 0.047 | 0.065 | -0.006 | 0.179 |
| Preserving Food | 0.043 | 0.052 | 0.412 | -0.059 | 0.145 |
| Foraging | 0.139 | 0.076 | 0.069 | -0.011 | 0.288 |
| Race/Ethnicity | -0.115 | 0.083 | 0.166 | -0.278 | 0.048 |
| Gender Identity | -0.213 | 0.046 | 0.000 | -0.304 | -0.123 |
| Age | 0.049 | 0.048 | 0.303 | -0.045 | 0.143 |
| Education Level | 0.057 | 0.045 | 0.206 | -0.031 | 0.145 |
| Household Income | 0.176 | 0.043 | 0.000 | 0.091 | 0.261 |
| Rurality | -0.053 | 0.043 | 0.220 | -0.138 | 0.032 |
| Job Disruption | 0.066 | 0.048 | 0.165 | -0.027 | 0.160 |
| Survey Year | 0.113 | 0.044 | 0.010 | 0.027 | 0.199 |
| Food Insecure |  |  |  |  |  |
| Gardening | 0.235 | 0.062 | 0.000 | 0.113 | 0.357 |
| Preserving Food | 0.009 | 0.061 | 0.882 | -0.111 | 0.129 |
| Foraging | -0.085 | 0.084 | 0.310 | -0.250 | 0.080 |
| Race/Ethnicity | -0.047 | 0.075 | 0.535 | -0.194 | 0.101 |
| Gender Identity | -0.347 | 0.064 | 0.000 | -0.473 | -0.221 |
| Age | 0.013 | 0.085 | 0.875 | -0.154 | 0.180 |
| Education Level | 0.056 | 0.061 | 0.359 | -0.064 | 0.177 |
| Household Income | 0.078 | 0.058 | 0.178 | -0.036 | 0.191 |
| Rurality | 0.095 | 0.056 | 0.092 | -0.015 | 0.204 |
| Job Disruption | -0.014 | 0.053 | 0.785 | -0.118 | 0.089 |
| Survey Year | -0.101 | 0.056 | 0.072 | -0.212 | 0.009 |

Statistically significant ( $p < 0.05$ ) results are bolded for emphasis.

### Hunting and Meat Consumption

Only 21.0% of respondents indicated they ate any game meat in the last 30 days, though 90.4% of respondents ate red meat and 94.1% ate white meat (Figure 2). Most (71.9%) respondents from households that hunted consumed game meat while only 13.5% of respondents from households that did not hunt consumed game meat. Sixteen percent of food secure respondents consumed game meat and 27.9% of food insecure respondents consumed game meat.

**Figure 2.**
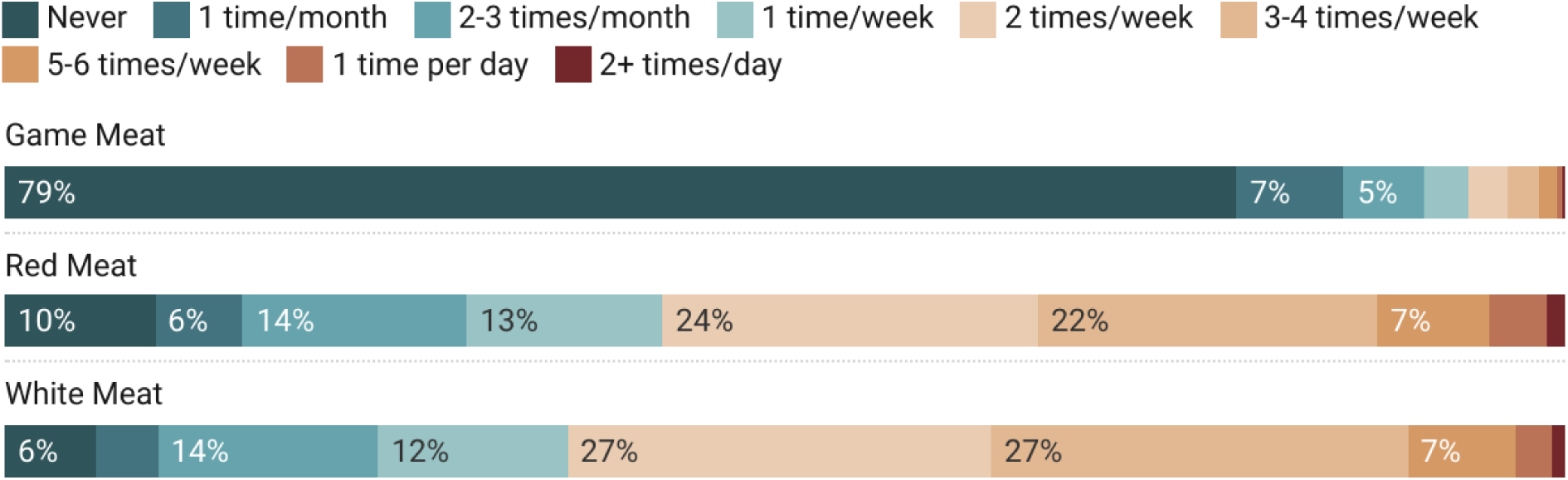
Consumption frequency of game meat, red meat, and white meat in the last 30 days.

Respondents whose households engaged in hunting were significantly more likely to consume game meat than respondents whose households did not hunt (H3a) (OR=17.25; p<0.001) (Table 9; full results of each model in supplementary material tables S25-S27). However, we also find that respondents who hunted were more likely to eat both red meat (OR=1.91; p<0.000) and white meat (OR=1.87; p<0.000) at greater frequencies than respondents who did not hunt (H3b). When looking at this relationship by food security status (H3c), both food secure (OR=23.72, p<0.001) and food insecure households that hunted (OR=13.27, p<0.001) were more likely to eat game meat than households that did not hunt (Table 10; full results of each model in supplementary material tables S28-S30). Likewise, both food secure (OR=1.99, p<0.001) and food insecure households that hunted (OR=2.23, p<0.001) were more likely to consume red meat in greater frequencies than households that did not hunt. Only food insecure households that hunted OR=1.75, p=0.006) were more likely to consume white meat in greater frequencies than food insecure households that did not hunt.

**Table 9.** Summary of results from separate logistic and ordinal logistic regression models predicting the effects of hunting on frequency of meat intake.

| **Dietary Factor** | **Odds Ratio** | **Standard Error** | **p-value** | **95% CI** |  |
| --- | --- | --- | --- | --- | --- |
| **Game Meat** | **17.25** | **3.050** | **0.000** | **12.193** | **24.390** |
| **Red Meat** | **1.91** | **0.227** | **0.000** | **1.513** | **2.411** |
| **White Meat** | **1.87** | **0.192** | **0.000** | **1.527** | **2.287** |

Each model also included control variables for age, gender identity, race/ethnicity, education level, income, rurality, job disruption, and survey year. Statistically significant ( $p < 0.05$ ) results are bolded for emphasis.

**Table 10.** Summary of results from separate logistic and ordinal logistic regression models predicting the effects of hunting on frequency of meat intake, by food security status.

| **Dietary Factor** | **Food Security Status** | **Odds Ratio** | **Standard Error** | **p-value** | **95% CI** |  |
| --- | --- | --- | --- | --- | --- | --- |
| Game Meat | **Food Secure** | **23.72** | **5.854** | **0.000** | **14.621** | **38.474** |
|  | **Food Insecure** | **13.27** | **3.656** | **0.000** | **7.731** | **22.767** |
| Red Meat | **Food Secure** | **1.99** | **0.327** | **0.000** | **1.446** | **2.749** |
|  | **Food Insecure** | **2.23** | **0.435** | **0.000** | **1.520** | **3.267** |
| White Meat | Food Secure | 1.25 | 0.183 | 0.134 | 0.935 | 1.660 |
|  | **Food Insecure** | **1.75** | **0.354** | **0.006** | **1.177** | **2.602** |

The model also included control variables for age, gender identity, race/ethnicity, education level, income, rurality, job disruption, and survey year. Statistically significant ( $p < 0.05$ ) results are bolded for emphasis.

## Discussion

This study, using a large-scale representative population sample from two rural U.S. states examined the relationship of a suite of HWFP activities to food security and dietary intake. It provides new evidence, with a large population size, of the relationship between HWFP engagement and food security status, fruit, vegetable, and meat intake, with notable implications for both individual and population-level health, as well as for HWFP as a strategy for affecting these outcomes.

Overall, our evidence suggests that several HWFP activities, including foraging, hunting, fishing, raising livestock, and raising poultry for eggs, were associated with greater odds of being food insecure, consistent with some of our previous findings ^(11)^. Food insecure households were more likely to engage in these activities than food secure households, perhaps as a coping strategy to supplement food from other sources. However, when combining all activities together into a single model, only households engaging in egg production were associated with greater odds of being food insecure.

In examining HWFP activities and their relationship to fruit and vegetable intake, we found that gardening, foraging, and preserving food are all positively and significantly associated with greater fruit and vegetable consumption. However, when combining all activities together into a single model, only gardening showed a significant association with greater fruit and vegetable consumption, with an approximately one cup-eq per week increase in consumption. Other studies have found that gardeners eat fruits and vegetables up to 2 more times per *day* than non-gardeners ^(18–20)^. One possible reason that we found a smaller difference in fruit and vegetable intake between respondents from gardening and non-gardening households, compared to what previous research has found, is that our survey’s recall period was just 30 days, and it was conducted in the springtime, before gardeners in Vermont and Maine were harvesting significant quantities of anything other than asparagus, rhubarb, and tender greens. Gardeners consistently self-report that the seasons when they harvest from their gardens coincide with eating greater quantities of vegetables ^(15,17,42)^. Given that the survey was conducted outside of harvest season, it is perhaps remarkable that we found a significant difference at all between gardeners’ and non-gardeners’ fruit and vegetable intakes and could be reflective of the high percentage of gardeners who also did food preservation or different food preferences between those who do and do not garden.

Furthermore, when exploring these relationships disaggregated by food security status, we find that this effect is stronger for food insecure households, especially in the combined model. As such, food insecure households engaging in gardening ate more than one and a half cup-eqs more fruit and vegetables per week, as compared to those not gardening. Food insecure households consume fewer fruits and vegetables overall compared to food secure households ^(43–45)^.

Furthermore, evidence early in the pandemic suggested that food insecure households were also more likely to reduce their fruit and vegetable intake ^(46,47)^. Food insecure individuals are also more likely to have a suite of other diet-related health challenges such as hypertension, diabetes, and cardiovascular disease ^(48,49)^, and increased fruit and vegetable intake may reduce disease risk or prevalence ^(50)^.

Our analysis also looked more closely at the relationship between hunting and wild game meat consumption, a relationship that has been seldom, if ever explored in population-level studies. We find an overall highly significant relationship between hunting and wild game consumption, which is especially true for food insecure respondents. Wild game meat can be an important source of protein and micronutrients and is typically lower in total and saturated fat than other sources of meat; thus, consuming game meat could have nutrition and health benefits ^(51)^.

However, in addition to being more likely to eat wild game meat, we also found that people who lived in households that engaged in hunting ate both red and white meat more frequently than people in non-hunting households. This suggests that hunting is not necessarily replacing other meat sources, but rather that it is being consumed in addition to red and white meat sources. This finding is consistent with previous research that showed people with connections to hunting or raising livestock reported more positive meat-related attitudes and more frequent meat consumption ^(52)^. Our findings are limited to examining meat intake frequency and future research should quantify the intake of different types of meat to examine whether people who hunt consume greater amounts of meat overall than people who don’t hunt.

Overall, this study offers one of the largest explorations of HWFP in a high-income country and its relationship to food security and dietary intake of fruits, vegetables, and wild game. Strengths of the study is the large sample size, representativeness, and inclusion of multiple HWFP activities. The majority of households that engaged in HFWP did multiple types of HWFP rather than just one activity, which highlights the importance of looking at a suite of activities. Several limitations should be noted, which we suggest can be the focus of future research. First, our findings only show correlations between HWFP activities and dietary outcomes. As noted, it may be that people who eat more fruits and vegetables are more likely to garden, rather than the other way around. Second, there was a mismatch between the period about which we asked about household HWFP engagement (the last year) and individual diet (the last 30 days). Further, there is a seasonal mismatch: the 30-day diet recall period did not fall during the half of the year when gardens are actively producing. The fact that members of gardening households still report eating more fruits and vegetables than non-gardeners could suggest early-season harvests or preserved bounty from the previous year, but it could also suggest that causation is in fact reversed, and households take up gardening because they eat more fruits and vegetables.

In reality, causation probably works in both directions to some extent, and only experimental research designs like that of Alaimo et al. (2023) can begin to disentangle gardening’s effect on fruit and vegetable consumption from fruit and vegetable consumption’s effect on gardening. Even in that case, truly randomized control trials are not possible with HWFP, since gardening, hunting, fishing, and foraging are not interventions that can be randomly applied across a population. It would be unethical to keep people who want to do these activities from engaging in them, and it is not plausible to force people who are not interested in these activities to do them—and nearly half of respondents who did not engage in HWFP reported that they are not interested ^(53)^. Even so, designing research that uses HWFP activities as an intervention may be a worthwhile direction for future research that can generate policy-relevant findings about the potential nutrition and health benefits of food self-provisioning.

## Conclusion

In this study, we show that HWFP engagement is associated with food security status. Respondents who engaged in these activities were more likely to be food insecure, indicating that food insecure households may be using HWFP as a coping strategy. We also find a positive association between HWFP engagement and fruit and vegetable intake, which may have important health implications, especially for food insecure individuals who typically have lower fruit and vegetable intakes than food secure individuals. Finally, we show a positive association between hunting and consumption of wild game meat. Our findings add to emerging evidence on the public health and food security benefits of HWFP engagement. We suggest that future research continues to examine these relationships over time to assess whether HWFP is an effective long-term strategy to improve health, well-being, and food security.

## Data Availability

All data produced in the present study are available upon reasonable request to the authors.

## Acknowledgements

We’d like to thank additional members of our teams in Vermont and Maine including Janica Anderzén, Jennifer Laurent, Jonathan Malacarne, Scott Merrill, Rebecca Mitchell, Sarah Nowak, Rachel Schattman, and Kate Yerxa.

## Funding Support

This work was supported by the following sources: a Joint Catalyst Award from the Gund Institute for Environment at the University of Vermont and the Northern New England Clinical and Translational Research Network; the USDA National Institute of Food and Agriculture (2022-67023-3645); University of Maine Agricultural and Forest Experiment Station (Hatch project numbers ME022103 and ME022122); and the UVM Food Systems Research Center via a Cooperative Agreement from the USDA Agricultural Research Service.

## Conflict of Interest

The authors declare no conflicts of interest.

## Authorship

ACM contributed to study design, data collection, analysis, writing, and revisions. AA contributed to study design, analysis, and writing. SB contributed to study design, data collection, writing, and revisions. FB, EHB, and KR contributed to study design, data collection, and revisions. MTN contributed to study design, data collection, writing, revisions, and project management.

## Ethical Standards Disclosure

This study was conducted according to the guidelines laid down in the Declaration of Helsinki and all procedures involving research study participants were approved by the University of Vermont Institutional Review Board (protocol no. 000000873) and the University of Maine Institutional Review Board (protocol no. 2020-07-11). Written informed consent was obtained from all subjects.

## Notes

### Competing Interest Statement

The authors have declared no competing interest.

### Author Declarations

This study was conducted according to the guidelines laid down in the Declaration of Helsinki and all procedures involving research study participants were approved by the University of Vermont Institutional Review Board (protocol no. 000000873) and the University of Maine Institutional Review Board (protocol no. 2020- 07-11). Written informed consent was obtained from all subjects.

