## Supplementary Materials for "Home and wild food procurement was associated with greater intake of fruits and vegetables during the COVID-19 pandemic in northern New England"

Table S1. Logistic regression results predicting the odds of food insecurity by any HWFP activity engagement. Odds ratios higher than 1.00 indicate a greater odds of food insecurity. Statistically significant (p<0.05) results are bolded for emphasis.

| | **Odds Ratio** | **Standard Error** | **p-value** | **95% CI** | |
|---|---|---|---|---|---|
| Any HWFP | 1.19 | 0.141 | 0.132 | 0.948 | 1.504 |
| Race/Ethnicity | 0.75 | 0.156 | 0.165 | 0.498 | 1.126 |
| Gender Identity | 1.00 | 0.124 | 0.996 | 0.784 | 1.277 |
| **Age** | **0.29** | **0.048** | **0.000** | **0.210** | **0.401** |
| **Education Level** | **0.51** | **0.061** | **0.000** | **0.400** | **0.640** |
| **Household Income** | **0.25** | **0.030** | **0.000** | **0.199** | **0.319** |
| Rurality | 1.02 | 0.117 | 0.845 | 0.817 | 1.280 |
| **Job Disruption** | **3.53** | **0.410** | **0.000** | **2.808** | **4.428** |
| **Survey Year** | **1.54** | **0.179** | **0.000** | **1.226** | **1.932** |

Table S2. Logistic regression results predicting the odds of food insecurity by gardening. Odds ratios higher than 1.00 indicate a greater odds of food insecurity. Statistically significant (p<0.05) results are bolded for emphasis.

| | **Odds Ratio** | **Standard Error** | **p-value** | **95% CI** | |
|---|---|---|---|---|---|
| Gardening | 1.06 | 0.123 | 0.606 | 0.846 | 1.331 |
| Race/Ethnicity | 0.75 | 0.157 | 0.173 | 0.501 | 1.133 |
| Gender Identity | 0.99 | 0.124 | 0.963 | 0.779 | 1.269 |
| **Age** | **0.29** | **0.048** | **0.000** | **0.209** | **0.400** |
| **Education Level** | **0.51** | **0.061** | **0.000** | **0.403** | **0.645** |
| **Household Income** | **0.26** | **0.031** | **0.000** | **0.201** | **0.323** |
| Rurality | 1.02 | 0.116 | 0.887 | 0.812 | 1.271 |
| **Job Disruption** | **3.54** | **0.411** | **0.000** | **2.823** | **4.449** |
| **Survey Year** | **1.56** | **0.180** | **0.000** | **1.240** | **1.951** |

Table S3. Logistic regression results predicting the odds of food insecurity by foraging. Odds ratios higher than 1.00 indicate a greater odds of food insecurity. Statistically significant ($p<0.05$) results are bolded for emphasis.

| | **Odds Ratio** | **Standard Error** | **p-value** | **95% CI** | |
|---|---|---|---|---|---|
| **Foraging** | **1.61** | **0.275** | **0.006** | **1.149** | **2.248** |
| Race/Ethnicity | 0.75 | 0.158 | 0.174 | 0.498 | 1.134 |
| Gender Identity | 1.04 | 0.131 | 0.750 | 0.813 | 1.332 |
| **Age** | **0.29** | **0.049** | **0.000** | **0.212** | **0.406** |
| **Education Level** | **0.51** | **0.061** | **0.000** | **0.401** | **0.642** |
| **Household Income** | **0.25** | **0.031** | **0.000** | **0.199** | **0.319** |
| Rurality | 1.04 | 0.120 | 0.730 | 0.831 | 1.303 |
| **Job Disruption** | **3.48** | **0.406** | **0.000** | **2.773** | **4.378** |
| **Survey Year** | **1.58** | **0.183** | **0.000** | **1.259** | **1.983** |

Table S4. Logistic regression results predicting the odds of food insecurity by hunting. Odds ratios higher than 1.00 indicate a greater odds of food insecurity. Statistically significant ($p<0.05$) results are bolded for emphasis.

| | **Odds Ratio** | **Standard Error** | **p-value** | **95% CI** | |
|---|---|---|---|---|---|
| **Hunting** | **1.64** | **0.277** | **0.003** | **1.177** | **2.283** |
| Race/Ethnicity | 0.75 | 0.156 | 0.170 | 0.501 | 1.130 |
| Gender Identity | 1.02 | 0.127 | 0.875 | 0.798 | 1.303 |
| **Age** | **0.30** | **0.049** | **0.000** | **0.216** | **0.412** |
| **Education Level** | **0.51** | **0.061** | **0.000** | **0.401** | **0.641** |
| **Household Income** | **0.25** | **0.030** | **0.000** | **0.199** | **0.319** |
| Rurality | 1.03 | 0.118 | 0.791 | 0.823 | 1.291 |
| **Job Disruption** | **3.58** | **0.416** | **0.000** | **2.847** | **4.491** |
| **Survey Year** | **1.60** | **0.185** | **0.000** | **1.273** | **2.004** |

Table S5. Logistic regression results predicting the odds of food insecurity by fishing. Odds ratios higher than 1.00 indicate a greater odds of food insecurity. Statistically significant ($p<0.05$) results are bolded for emphasis.

| | Odds Ratio | Standard Error | p-value | 95% CI | |
|---|---|---|---|---|---|
| **Fishing** | **1.60** | **0.264** | **0.005** | **1.156** | **2.208** |
| Race/Ethnicity | 0.76 | 0.160 | 0.195 | 0.504 | 1.151 |
| Gender Identity | 1.04 | 0.129 | 0.765 | 0.813 | 1.325 |
| **Age** | **0.30** | **0.049** | **0.000** | **0.218** | **0.414** |
| **Education Level** | **0.51** | **0.061** | **0.000** | **0.400** | **0.640** |
| **Household Income** | **0.25** | **0.031** | **0.000** | **0.200** | **0.320** |
| Rurality | 1.04 | 0.120 | 0.710 | 0.833 | 1.307 |
| **Job Disruption** | **3.53** | **0.411** | **0.000** | **2.809** | **4.436** |
| **Survey Year** | **1.56** | **0.180** | **0.000** | **1.240** | **1.952** |

Table S6. Logistic regression results predicting the odds of food insecurity by preserving food. Odds ratios higher than 1.00 indicate a greater odds of food insecurity. Statistically significant ($p<0.05$) results are bolded for emphasis.

| | Odds Ratio | Standard Error | p-value | 95% CI | |
|---|---|---|---|---|---|
| Preserving Food | 1.22 | 0.159 | 0.128 | 0.945 | 1.574 |
| Race/Ethnicity | 0.75 | 0.156 | 0.168 | 0.499 | 1.129 |
| Gender Identity | 0.99 | 0.124 | 0.954 | 0.778 | 1.267 |
| **Age** | **0.29** | **0.047** | **0.000** | **0.209** | **0.398** |
| **Education Level** | **0.50** | **0.061** | **0.000** | **0.397** | **0.637** |
| **Household Income** | **0.26** | **0.031** | **0.000** | **0.202** | **0.323** |
| Rurality | 1.01 | 0.116 | 0.908 | 0.809 | 1.269 |
| **Job Disruption** | **3.52** | **0.409** | **0.000** | **2.798** | **4.417** |
| **Survey Year** | **1.53** | **0.179** | **0.000** | **1.213** | **1.920** |

Table S7. Logistic regression results predicting the odds of food insecurity by raising livestock. Odds ratios higher than 1.00 indicate a greater odds of food insecurity. Statistically significant ($p<0.05$) results are bolded for emphasis.

| | Odds Ratio | Standard Error | p-value | 95% CI | |
|---|---|---|---|---|---|
| **Raising Livestock** | **2.71** | **0.733** | **0.000** | **1.592** | **4.602** |
| Race/Ethnicity | 0.74 | 0.153 | 0.142 | 0.491 | 1.108 |
| Gender Identity | 1.04 | 0.130 | 0.751 | 0.815 | 1.328 |
| **Age** | **0.30** | **0.050** | **0.000** | **0.218** | **0.417** |
| **Education Level** | **0.51** | **0.061** | **0.000** | **0.406** | **0.648** |
| **Household Income** | **0.26** | **0.031** | **0.000** | **0.202** | **0.325** |
| Rurality | 1.02 | 0.117 | 0.856 | 0.816 | 1.278 |
| **Job Disruption** | **3.60** | **0.419** | **0.000** | **2.865** | **4.523** |
| **Survey Year** | **1.65** | **0.192** | **0.000** | **1.310** | **2.069** |

Table S8. Logistic regression results predicting the odds of food insecurity by raising poultry for eggs. Odds ratios higher than 1.00 indicate a greater odds of food insecurity. Statistically significant ($p<0.05$) results are bolded for emphasis.

| | Odds Ratio | Standard Error | p-value | 95% CI | |
|---|---|---|---|---|---|
| **Raising Poultry for Eggs** | **2.07** | **0.390** | **0.000** | **1.433** | **2.997** |
| Race/Ethnicity | 0.75 | 0.154 | 0.163 | 0.502 | 1.123 |
| Gender Identity | 1.01 | 0.125 | 0.906 | 0.796 | 1.293 |
| **Age** | **0.30** | **0.050** | **0.000** | **0.220** | **0.420** |
| **Education Level** | **0.52** | **0.062** | **0.000** | **0.409** | **0.654** |
| **Household Income** | **0.25** | **0.031** | **0.000** | **0.199** | **0.320** |
| Rurality | 1.03 | 0.118 | 0.803 | 0.822 | 1.288 |
| **Job Disruption** | **3.53** | **0.411** | **0.000** | **2.813** | **4.438** |
| **Survey Year** | **1.58** | **0.183** | **0.000** | **1.259** | **1.983** |

Table S9. Linear regression results predicting the effects of gardening on daily fruit and vegetable intake (cup equivalents). Statistically significant ($p<0.05$) results are bolded for emphasis.

| Fruit and Vegetable Intake | Coefficient | Standard Error | p-value | 95% CI | |
|---|---|---|---|---|---|
| **Gardening** | **0.152** | **0.033** | **0.000** | **0.087** | **0.217** |
| Race/Ethnicity | -0.044 | 0.058 | 0.450 | -0.157 | 0.070 |
| **Gender Identity** | **-0.254** | **0.036** | **0.000** | **-0.325** | **-0.184** |
| **Age** | **0.085** | **0.039** | **0.030** | **0.008** | **0.162** |
| **Education Level** | **0.089** | **0.034** | **0.010** | **0.021** | **0.156** |
| **Household Income** | **0.194** | **0.034** | **0.000** | **0.128** | **0.260** |
| Rurality | 0.009 | 0.034 | 0.779 | -0.057 | 0.076 |
| Job Disruption | -0.004 | 0.035 | 0.901 | -0.073 | 0.064 |
| Survey Year | 0.033 | 0.032 | 0.304 | -0.030 | 0.097 |

Table S10. Linear regression results predicting the effects of gardening on daily fruit intake (cup equivalents). Statistically significant ($p<0.05$) results are bolded for emphasis.

| Fruit Intake | Coefficient | Standard Error | p-value | 95% CI | |
|---|---|---|---|---|---|
| **Gardening** | **0.068** | **0.019** | **0.000** | **0.031** | **0.104** |
| Race/Ethnicity | -0.051 | 0.038 | 0.175 | -0.124 | 0.023 |
| **Gender Identity** | **-0.053** | **0.021** | **0.011** | **-0.094** | **-0.012** |
| Age | -0.002 | 0.021 | 0.919 | -0.044 | 0.040 |
| **Education Level** | **0.038** | **0.019** | **0.049** | **0.000** | **0.075** |
| **Household Income** | **0.055** | **0.019** | **0.004** | **0.018** | **0.092** |
| Rurality | 0.002 | 0.019 | 0.907 | -0.035 | 0.040 |
| Job Disruption | -0.005 | 0.020 | 0.793 | -0.044 | 0.034 |
| Survey Year | 0.019 | 0.018 | 0.305 | -0.017 | 0.054 |

Table S11. Linear regression results predicting the effects of gardening on daily vegetable intake (cup equivalents). Statistically significant ($p<0.05$) results are bolded for emphasis.

| Vegetable Intake | Coefficient | Standard Error | p-value | 95% CI | |
|---|---|---|---|---|---|
| **Gardening** | **0.088** | **0.020** | **0.000** | **0.049** | **0.127** |
| Race/Ethnicity | -0.017 | 0.034 | 0.612 | -0.084 | 0.049 |
| **Gender Identity** | **-0.209** | **0.022** | **0.000** | **-0.252** | **-0.167** |
| **Age** | **0.082** | **0.025** | **0.001** | **0.033** | **0.131** |
| Education Level | 0.036 | 0.021 | 0.085 | -0.005 | 0.077 |
| **Household Income** | **0.120** | **0.020** | **0.000** | **0.081** | **0.160** |
| Rurality | 0.011 | 0.021 | 0.581 | -0.029 | 0.052 |
| Job Disruption | 0.012 | 0.021 | 0.576 | -0.030 | 0.053 |
| Survey Year | 0.016 | 0.020 | 0.410 | -0.022 | 0.055 |

Table S12. Linear regression results predicting the effects of preserving food on daily fruit and vegetable intake (cup equivalents). Statistically significant ($p<0.05$) results are bolded for emphasis.

| Fruit and Vegetable Intake | Coefficient | Standard Error | p-value | 95% CI | |
|---|---|---|---|---|---|
| **Preservation** | **0.087** | **0.037** | **0.019** | **0.014** | **0.160** |
| Race/Ethnicity | -0.048 | 0.058 | 0.408 | -0.162 | 0.066 |
| **Gender Identity** | **-0.256** | **0.036** | **0.000** | **-0.328** | **-0.185** |
| **Age** | **0.091** | **0.040** | **0.022** | **0.013** | **0.169** |
| **Education Level** | **0.094** | **0.035** | **0.007** | **0.025** | **0.163** |
| **Household Income** | **0.207** | **0.034** | **0.000** | **0.142** | **0.273** |
| Rurality | 0.002 | 0.034 | 0.962 | -0.065 | 0.068 |
| Job Disruption | -0.004 | 0.035 | 0.912 | -0.073 | 0.066 |
| Survey Year | 0.029 | 0.033 | 0.379 | -0.036 | 0.094 |

Table S13. Linear regression results predicting the effects of preserving food on daily fruit intake (cup equivalents). Statistically significant ($p<0.05$) results are bolded for emphasis.

| Fruit Intake | Coefficient | Standard Error | p-value | 95% CI | |
|---|---|---|---|---|---|
| **Preservation** | **0.051** | **0.021** | **0.018** | **0.009** | **0.093** |
| Race/Ethnicity | -0.053 | 0.038 | 0.158 | -0.127 | 0.021 |
| **Gender Identity** | **-0.054** | **0.021** | **0.011** | **-0.096** | **-0.012** |
| Age | 0.000 | 0.021 | 0.993 | -0.042 | 0.042 |
| **Education Level** | **0.039** | **0.019** | **0.044** | **0.001** | **0.077** |
| **Household Income** | **0.061** | **0.019** | **0.001** | **0.024** | **0.098** |
| Rurality | -0.001 | 0.019 | 0.951 | -0.039 | 0.036 |
| Job Disruption | -0.006 | 0.020 | 0.767 | -0.045 | 0.034 |
| Survey Year | 0.015 | 0.019 | 0.409 | -0.021 | 0.052 |

Table S14. Linear regression results predicting the effects of preserving food on daily vegetable intake (cup equivalents). Statistically significant ($p<0.05$) results are bolded for emphasis.

| Vegetable Intake | Coefficient | Standard Error | p-value | 95% CI | |
|---|---|---|---|---|---|
| **Preservation** | **0.049** | **0.022** | **0.028** | **0.005** | **0.093** |
| Race/Ethnicity | -0.020 | 0.034 | 0.562 | -0.086 | 0.047 |
| **Gender Identity** | **-0.210** | **0.022** | **0.000** | **-0.253** | **-0.167** |
| **Age** | **0.085** | **0.025** | **0.001** | **0.036** | **0.134** |
| Education Level | 0.039 | 0.021 | 0.065 | -0.002 | 0.081 |
| **Household Income** | **0.128** | **0.020** | **0.000** | **0.088** | **0.168** |
| Rurality | 0.007 | 0.021 | 0.740 | -0.033 | 0.047 |
| Job Disruption | 0.012 | 0.021 | 0.570 | -0.030 | 0.054 |
| Survey Year | 0.014 | 0.020 | 0.485 | -0.025 | 0.053 |

Table S15. Linear regression results predicting the effects of foraging on daily fruit and vegetable intake (cup equivalents). Statistically significant ($p<0.05$) results are bolded for emphasis.

| **Fruit and Vegetable Intake** | **Coefficient** | **Standard Error** | **p-value** | **95% CI** | |
|---|---|---|---|---|---|
| **Foraging** | **0.106** | **0.051** | **0.038** | **0.006** | **0.206** |
| Race/Ethnicity | -0.045 | 0.058 | 0.444 | -0.159 | 0.070 |
| **Gender Identity** | **-0.247** | **0.037** | **0.000** | **-0.319** | **-0.174** |
| **Age** | **0.101** | **0.040** | **0.012** | **0.023** | **0.179** |
| **Education Level** | **0.100** | **0.035** | **0.004** | **0.031** | **0.168** |
| **Household Income** | **0.206** | **0.034** | **0.000** | **0.140** | **0.272** |
| Rurality | 0.006 | 0.034 | 0.858 | -0.060 | 0.073 |
| Job Disruption | -0.002 | 0.036 | 0.946 | -0.072 | 0.067 |
| Survey Year | 0.042 | 0.033 | 0.202 | -0.022 | 0.105 |

Table S16. Linear regression results predicting the effects of foraging on daily fruit intake (cup equivalents). Statistically significant ($p<0.05$) results are bolded for emphasis.

| **Fruit Intake** | **Coefficient** | **Standard Error** | **p-value** | **95% CI** | |
|---|---|---|---|---|---|
| Foraging | 0.033 | 0.029 | 0.248 | -0.023 | 0.089 |
| Race/Ethnicity | -0.051 | 0.038 | 0.175 | -0.125 | 0.023 |
| **Gender Identity** | **-0.051** | **0.022** | **0.018** | **-0.093** | **-0.009** |
| Age | 0.004 | 0.022 | 0.842 | -0.038 | 0.047 |
| **Education Level** | **0.043** | **0.019** | **0.027** | **0.005** | **0.080** |
| **Household Income** | **0.061** | **0.019** | **0.001** | **0.024** | **0.098** |
| Rurality | 0.000 | 0.019 | 0.995 | -0.037 | 0.038 |
| Job Disruption | -0.004 | 0.020 | 0.858 | -0.043 | 0.036 |
| Survey Year | 0.022 | 0.018 | 0.226 | -0.014 | 0.058 |

Table S17. Linear regression results predicting the effects of foraging on daily vegetable intake (cup equivalents). Statistically significant (p<0.05) results are bolded for emphasis.

| **Vegetable Intake** | **Coefficient** | **Standard Error** | **p-value** | **95% CI** | |
|---|---|---|---|---|---|
| **Foraging** | **0.078** | **0.031** | **0.012** | **0.017** | **0.138** |
| Race/Ethnicity | -0.018 | 0.034 | 0.601 | -0.084 | 0.049 |
| **Gender Identity** | **-0.203** | **0.022** | **0.000** | **-0.247** | **-0.160** |
| **Age** | **0.092** | **0.025** | **0.000** | **0.043** | **0.141** |
| **Education Level** | **0.042** | **0.021** | **0.045** | **0.001** | **0.084** |
| **Household Income** | **0.127** | **0.020** | **0.000** | **0.087** | **0.167** |
| Rurality | 0.010 | 0.021 | 0.622 | -0.030 | 0.050 |
| Job Disruption | 0.012 | 0.022 | 0.575 | -0.030 | 0.054 |
| Survey Year | 0.021 | 0.020 | 0.281 | -0.017 | 0.060 |

Table S18. Linear regression results predicting the combined effects of gardening, foraging, and preserving food on daily fruit and vegetable intake (cup equivalents). Statistically significant (p<0.05) results are bolded for emphasis.

| **Fruit and Vegetable Intake** | **Coefficient** | **Standard Error** | **p-value** | **95% CI** | |
|---|---|---|---|---|---|
| **Gardening** | **0.135** | **0.037** | **0.000** | **0.064** | **0.207** |
| Preserving Food | 0.028 | 0.040 | 0.487 | -0.050 | 0.106 |
| Foraging | 0.045 | 0.056 | 0.421 | -0.064 | 0.154 |
| Race/Ethnicity | -0.045 | 0.058 | 0.438 | -0.159 | 0.069 |
| **Gender Identity** | **-0.251** | **0.037** | **0.000** | **-0.323** | **-0.179** |
| **Age** | **0.087** | **0.040** | **0.029** | **0.009** | **0.165** |
| **Education Level** | **0.087** | **0.035** | **0.012** | **0.019** | **0.155** |
| **Household Income** | **0.194** | **0.033** | **0.000** | **0.128** | **0.260** |
| Rurality | 0.011 | 0.034 | 0.750 | -0.055 | 0.077 |
| Job Disruption | -0.008 | 0.035 | 0.818 | -0.078 | 0.061 |
| Survey Year | 0.031 | 0.033 | 0.343 | -0.034 | 0.096 |

Table S19. Linear regression results predicting the combined effects of gardening, foraging, and preserving food on daily fruit intake (cup equivalents). Statistically significant ($p<0.05$) results are bolded for emphasis.

| **Fruit and Vegetable Intake** | **Coefficient** | **Standard Error** | **p-value** | **95% CI** | |
|---|---|---|---|---|---|
| **Gardening** | **0.058** | **0.021** | **0.005** | **0.018** | **0.099** |
| Preserving Food | 0.029 | 0.023 | 0.212 | -0.016 | 0.074 |
| Foraging | 0.001 | 0.030 | 0.981 | -0.059 | 0.060 |
| Race/Ethnicity | -0.052 | 0.038 | 0.166 | -0.126 | 0.022 |
| **Gender Identity** | **-0.053** | **0.021** | **0.013** | **-0.095** | **-0.011** |
| Age | -0.003 | 0.022 | 0.888 | -0.045 | 0.039 |
| Education Level | 0.036 | 0.019 | 0.060 | -0.002 | 0.074 |
| **Household Income** | **0.055** | **0.019** | **0.004** | **0.018** | **0.093** |
| Rurality | 0.002 | 0.019 | 0.921 | -0.036 | 0.039 |
| Job Disruption | -0.007 | 0.020 | 0.722 | -0.046 | 0.032 |
| Survey Year | 0.016 | 0.019 | 0.403 | -0.021 | 0.052 |

Table S20. Linear regression results predicting the combined effects of gardening, foraging, and preserving food on daily vegetable intake (cup equivalents). Statistically significant ($p<0.05$) results are bolded for emphasis.

| **Fruit and Vegetable Intake** | **Coefficient** | **Standard Error** | **p-value** | **95% CI** | |
|---|---|---|---|---|---|
| Gardening | 0.077 | 0.022 | 3.530 | 0.034 | 0.120 |
| Preserving Food | 0.012 | 0.024 | 0.627 | -0.035 | 0.059 |
| Foraging | 0.044 | 0.034 | 0.190 | -0.022 | 0.111 |
| Race/Ethnicity | -0.018 | 0.034 | 0.601 | -0.084 | 0.049 |
| **Gender Identity** | **-0.206** | **0.022** | **0.000** | **-0.249** | **-0.162** |
| **Age** | **0.084** | **0.025** | **0.001** | **0.035** | **0.133** |
| Education Level | 0.036 | 0.021 | 0.090 | -0.006 | 0.077 |
| **Household Income** | **0.120** | **0.020** | **0.000** | **0.080** | **0.160** |
| Rurality | 0.013 | 0.021 | 0.531 | -0.027 | 0.053 |
| Job Disruption | 0.009 | 0.022 | 0.673 | -0.033 | 0.051 |
| Survey Year | 0.016 | 0.020 | 0.423 | -0.023 | 0.055 |

Table S21. Linear regression results predicting the effects of gardening on daily combined fruit and vegetable intake (cup equivalents), by food security status. Statistically significant ($p<0.05$) results are bolded for emphasis.

| **Fruit and Vegetable Intake** | **Coefficient** | **Standard Error** | **p-value** | **95% CI** | |
|---|---|---|---|---|---|
| | | Food Secure | | | |
| **Gardening** | **0.120** | **0.043** | **0.006** | **0.035** | **0.205** |
| Race/Ethnicity | -0.114 | 0.084 | 0.175 | -0.279 | 0.051 |
| **Gender Identity** | **-0.222** | **0.045** | **0.000** | **-0.311** | **-0.132** |
| Age | 0.044 | 0.047 | 0.351 | -0.049 | 0.137 |
| Education Level | 0.057 | 0.045 | 0.203 | -0.031 | 0.146 |
| **Household Income** | **0.177** | **0.044** | **0.000** | **0.092** | **0.263** |
| Rurality | -0.060 | 0.043 | 0.165 | -0.144 | 0.025 |
| Job Disruption | 0.081 | 0.047 | 0.087 | -0.012 | 0.173 |
| **Survey Year** | **0.120** | **0.043** | **0.005** | **0.037** | **0.204** |
| | | Food Insecure | | | |
| **Gardening** | **0.220** | **0.055** | **0.000** | **0.113** | **0.327** |
| Race/Ethnicity | -0.049 | 0.076 | 0.519 | -0.197 | 0.100 |
| **Gender Identity** | **-0.337** | **0.063** | **0.000** | **-0.461** | **-0.212** |
| Age | 0.018 | 0.085 | 0.830 | -0.149 | 0.185 |
| Education Level | 0.054 | 0.061 | 0.372 | -0.065 | 0.174 |
| Household Income | 0.077 | 0.057 | 0.179 | -0.035 | 0.189 |
| Rurality | 0.099 | 0.057 | 0.081 | -0.012 | 0.210 |
| Job Disruption | -0.015 | 0.053 | 0.781 | -0.119 | 0.090 |
| Survey Year | -0.096 | 0.057 | 0.092 | -0.208 | 0.016 |

Table S22. Linear regression results predicting the effects of preserving food on daily combined fruit and vegetable intake (cup equivalents), by food security status. Statistically significant (p<0.05) results are bolded for emphasis.

| **Fruit and Vegetable Intake** | **Coefficient** | **Standard Error** | **p-value** | **95% CI** | |
|---|---|---|---|---|---|
| | | Food Secure | | | |
| **Preservation** | **0.096** | **0.049** | **0.050** | **0.000** | **0.192** |
| Race/Ethnicity | -0.123 | 0.084 | 0.141 | -0.287 | 0.041 |
| **Gender Identity** | **-0.221** | **0.046** | **0.000** | **-0.311** | **-0.131** |
| Age | 0.051 | 0.047 | 0.279 | -0.041 | 0.143 |
| Education Level | 0.062 | 0.045 | 0.173 | -0.027 | 0.151 |
| **Household Income** | **0.186** | **0.043** | **0.000** | **0.101** | **0.271** |
| Rurality | -0.062 | 0.043 | 0.151 | -0.147 | 0.023 |
| Job Disruption | 0.075 | 0.047 | 0.114 | -0.018 | 0.168 |
| **Survey Year** | **0.109** | **0.044** | **0.013** | **0.023** | **0.195** |
| | | Food Insecure | | | |
| Preservation | 0.070 | 0.060 | 0.245 | -0.048 | 0.188 |
| Race/Ethnicity | -0.039 | 0.076 | 0.603 | -0.189 | 0.110 |
| **Gender Identity** | **-0.348** | **0.065** | **0.000** | **-0.476** | **-0.220** |
| Age | -0.009 | 0.087 | 0.919 | -0.179 | 0.161 |
| Education Level | 0.062 | 0.063 | 0.326 | -0.062 | 0.185 |
| Household Income | 0.103 | 0.060 | 0.088 | -0.015 | 0.221 |
| Rurality | 0.082 | 0.056 | 0.144 | -0.028 | 0.192 |
| Job Disruption | -0.004 | 0.054 | 0.947 | -0.110 | 0.103 |
| Survey Year | -0.095 | 0.058 | 0.101 | -0.208 | 0.018 |

Table S23. Linear regression results predicting the effects of foraging on daily combined fruit and vegetable intake (cup equivalents), by food security status. Statistically significant ($p<0.05$) results are bolded for emphasis.

| **Fruit and Vegetable Intake** | **Coefficient** | **Standard Error** | **p-value** | **95% CI** | |
|---|---|---|---|---|---|
| | | Food Secure | | | |
| **Foraging** | **0.182** | **0.070** | **0.009** | **0.045** | **0.319** |
| Race/Ethnicity | -0.118 | 0.084 | 0.159 | -0.283 | 0.047 |
| **Gender Identity** | **-0.209** | **0.046** | **0.000** | **-0.300** | **-0.119** |
| Age | 0.064 | 0.047 | 0.173 | -0.028 | 0.157 |
| Education Level | 0.069 | 0.045 | 0.122 | -0.019 | 0.157 |
| **Household Income** | **0.180** | **0.043** | **0.000** | **0.094** | **0.265** |
| Rurality | -0.056 | 0.043 | 0.198 | -0.141 | 0.029 |
| Job Disruption | 0.074 | 0.048 | 0.123 | -0.020 | 0.168 |
| **Survey Year** | **0.125** | **0.042** | **0.003** | **0.042** | **0.209** |
| | | Food Insecure | | | |
| Foraging | 0.015 | 0.080 | 0.851 | -0.142 | 0.172 |
| Race/Ethnicity | -0.037 | 0.075 | 0.620 | -0.185 | 0.111 |
| **Gender Identity** | **-0.342** | **0.067** | **0.000** | **-0.474** | **-0.210** |
| Age | -0.008 | 0.088 | 0.925 | -0.182 | 0.165 |
| Education Level | 0.067 | 0.063 | 0.284 | -0.056 | 0.190 |
| Household Income | 0.107 | 0.061 | 0.078 | -0.012 | 0.226 |
| Rurality | 0.086 | 0.057 | 0.130 | -0.025 | 0.197 |
| Job Disruption | -0.006 | 0.054 | 0.919 | -0.112 | 0.101 |
| Survey Year | -0.094 | 0.057 | 0.103 | -0.206 | 0.019 |

Table S24. Linear regression results predicting the effects of engagement in any HWFP activity on daily combined fruit and vegetable intake (cup equivalents), by food security status. Statistically significant (p<0.05) results are bolded for emphasis.

| **Fruit and Vegetable Intake** | **Coefficient** | **Standard Error** | **p-value** | **95% CI** | |
|---|---|---|---|---|---|
| | | Food Secure | | | |
| Any HWFP | 0.079 | 0.044 | 0.074 | -0.008 | 0.165 |
| Race/Ethnicity | -0.123 | 0.085 | 0.147 | -0.289 | 0.043 |
| **Gender Identity** | **-0.222** | **0.046** | **0.000** | **-0.312** | **-0.133** |
| Age | 0.055 | 0.047 | 0.239 | -0.037 | 0.147 |
| Education Level | 0.062 | 0.045 | 0.173 | -0.027 | 0.150 |
| **Household Income** | **0.181** | **0.044** | **0.000** | **0.095** | **0.267** |
| Rurality | -0.059 | 0.043 | 0.171 | -0.145 | 0.026 |
| Job Disruption | 0.084 | 0.047 | 0.077 | -0.009 | 0.177 |
| **Survey Year** | **0.119** | **0.043** | **0.005** | **0.035** | **0.202** |
| | | Food Insecure | | | |
| **Any HWFP** | **0.140** | **0.050** | **0.005** | **0.041** | **0.238** |
| Race/Ethnicity | -0.042 | 0.076 | 0.584 | -0.192 | 0.108 |
| **Gender Identity** | **-0.337** | **0.064** | **0.000** | **-0.463** | **-0.211** |
| Age | 0.014 | 0.088 | 0.872 | -0.158 | 0.186 |
| Education Level | 0.063 | 0.062 | 0.309 | -0.059 | 0.186 |
| Household Income | 0.086 | 0.059 | 0.144 | -0.029 | 0.201 |
| Rurality | 0.089 | 0.056 | 0.116 | -0.022 | 0.200 |
| Job Disruption | -0.010 | 0.054 | 0.857 | -0.115 | 0.096 |
| Survey Year | -0.101 | 0.057 | 0.077 | -0.214 | 0.011 |

Table S25. Logistic regression results predicting the effects of hunting on frequency of game meat intake. Statistically significant (p<0.05) results are bolded for emphasis.

| **Game Meat Intake Frequency** | **Odds Ratio** | **Standard Error** | **p-value** | **95% CI** | |
|---|---|---|---|---|---|
| **Hunting** | **17.25** | **3.050** | **0.000** | **12.193** | **24.390** |
| Race/Ethnicity | 0.87 | 0.231 | 0.612 | 0.521 | 1.469 |
| **Gender Identity** | **0.52** | **0.074** | **0.000** | **0.391** | **0.683** |
| **Age** | **0.33** | **0.063** | **0.000** | **0.228** | **0.482** |
| Education Level | 0.84 | 0.117 | 0.201 | 0.635 | 1.100 |
| Household Income | 0.94 | 0.133 | 0.666 | 0.713 | 1.242 |
| Rurality | 0.99 | 0.138 | 0.961 | 0.757 | 1.303 |
| **Job Disruption** | **1.43** | **0.196** | **0.010** | **1.089** | **1.867** |
| Survey Year | 1.16 | 0.157 | 0.279 | 0.888 | 1.512 |

Table S26. Ordinal logistic regression results predicting the effects of hunting on frequency of red meat intake. Statistically significant ($p<0.05$) results are bolded for emphasis.

| **Red Meat Intake Frequency** | **Odds Ratio** | **Standard Error** | **p-value** | **95% CI** | |
|---|---|---|---|---|---|
| **Hunting** | **1.91** | **0.227** | **0.000** | **1.513** | **2.411** |
| Race/Ethnicity | 0.97 | 0.154 | 0.868 | 0.714 | 1.329 |
| **Gender Identity** | **0.70** | **0.062** | **0.000** | **0.586** | **0.832** |
| Age | 1.04 | 0.106 | 0.693 | 0.852 | 1.272 |
| **Education Level** | **0.80** | **0.070** | **0.011** | **0.675** | **0.951** |
| Household Income | 1.17 | 0.105 | 0.073 | 0.985 | 1.399 |
| Rurality | 0.91 | 0.077 | 0.246 | 0.767 | 1.070 |
| Job Disruption | 0.94 | 0.086 | 0.504 | 0.787 | 1.125 |
| Survey Year | 0.98 | 0.082 | 0.832 | 0.835 | 1.157 |

Table S27. Ordinal logistic regression results predicting the effects of hunting on frequency of white meat intake. Statistically significant ($p<0.05$) results are bolded for emphasis.

| **White Meat Intake Frequency** | **Odds Ratio** | **Standard Error** | **p-value** | **95% CI** | |
|---|---|---|---|---|---|
| **Hunting** | **1.36** | **0.159** | **0.009** | **1.080** | **1.710** |
| Race/Ethnicity | 0.79 | 0.138 | 0.171 | 0.558 | 1.109 |
| Gender Identity | 1.08 | 0.095 | 0.361 | 0.912 | 1.288 |
| **Age** | **0.78** | **0.074** | **0.010** | **0.653** | **0.944** |
| Education Level | 0.95 | 0.084 | 0.561 | 0.799 | 1.129 |
| **Household Income** | **1.74** | **0.157** | **0.000** | **1.456** | **2.075** |
| Rurality | 1.16 | 0.100 | 0.081 | 0.982 | 1.375 |
| **Job Disruption** | **1.25** | **0.115** | **0.016** | **1.043** | **1.497** |
| Survey Year | 0.96 | 0.081 | 0.621 | 0.813 | 1.132 |

Table S28. Logistic regression results predicting the effects of hunting on game meat intake, by food security status. Statistically significant (p<0.05) results are bolded for emphasis.

| **Game Meat Intake Frequency** | **Odds Ratio** | **Standard Error** | **p-value** | **95% CI** | |
|---|---|---|---|---|---|
| | | Food Secure | | | |
| **Hunting** | **23.72** | **5.854** | **0.000** | **14.621** | **38.474** |
| Race/Ethnicity | 1.17 | 0.553 | 0.738 | 0.464 | 2.954 |
| **Gender Identity** | **0.56** | **0.120** | **0.007** | **0.368** | **0.852** |
| **Age** | **0.42** | **0.101** | **0.000** | **0.265** | **0.676** |
| Education Level | 0.83 | 0.172 | 0.369 | 0.554 | 1.245 |
| Household Income | 1.21 | 0.260 | 0.381 | 0.792 | 1.841 |
| Rurality | 0.87 | 0.178 | 0.486 | 0.579 | 1.297 |
| Job Disruption | 1.16 | 0.261 | 0.521 | 0.742 | 1.800 |
| Survey Year | 1.28 | 0.265 | 0.233 | 0.853 | 1.921 |
| | | Food Insecure | | | |
| **Hunting** | **13.27** | **3.656** | **0.000** | **7.731** | **22.767** |
| Race/Ethnicity | 0.95 | 0.348 | 0.896 | 0.466 | 1.949 |
| **Gender Identity** | **0.43** | **0.091** | **0.000** | **0.284** | **0.650** |
| **Age** | **0.28** | **0.119** | **0.003** | **0.121** | **0.644** |
| Education Level | 1.15 | 0.244 | 0.515 | 0.757 | 1.743 |
| Household Income | 1.09 | 0.238 | 0.700 | 0.708 | 1.671 |
| Rurality | 1.11 | 0.230 | 0.619 | 0.738 | 1.666 |
| Job Disruption | 1.23 | 0.256 | 0.316 | 0.819 | 1.852 |
| Survey Year | 0.98 | 0.200 | 0.920 | 0.657 | 1.461 |

Table S29. Ordinal logistic regression results predicting the effects of hunting on frequency of red meat intake, by food security status. Statistically significant ($p<0.05$) results are bolded for emphasis.

| **Red Meat Intake Frequency** | **Odds Ratio** | **Standard Error** | **p-value** | **95% CI** | |
|---|---|---|---|---|---|
| | | Food Secure | | | |
| **Hunting** | **1.99** | **0.327** | **0.000** | **1.446** | **2.749** |
| Race/Ethnicity | 1.12 | 0.296 | 0.663 | 0.669 | 1.880 |
| **Gender Identity** | **0.63** | **0.074** | **0.000** | **0.500** | **0.791** |
| Age | 1.02 | 0.128 | 0.849 | 0.801 | 1.309 |
| **Education Level** | **0.73** | **0.085** | **0.006** | **0.580** | **0.915** |
| Household Income | 1.16 | 0.141 | 0.235 | 0.910 | 1.469 |
| **Rurality** | **0.75** | **0.083** | **0.009** | **0.601** | **0.929** |
| Job Disruption | 0.97 | 0.130 | 0.820 | 0.746 | 1.261 |
| Survey Year | 0.90 | 0.100 | 0.363 | 0.729 | 1.123 |
| | | Food Insecure | | | |
| Hunting | 2.23 | 0.435 | 4.110 | 1.520 | 3.267 |
| Race/Ethnicity | 0.79 | 0.162 | 0.244 | 0.525 | 1.178 |
| Gender Identity | 0.85 | 0.132 | 0.303 | 0.631 | 1.154 |
| Age | 0.74 | 0.182 | 0.222 | 0.457 | 1.199 |
| Education Level | 0.84 | 0.129 | 0.251 | 0.620 | 1.133 |
| Household Income | 0.97 | 0.151 | 0.846 | 0.715 | 1.317 |
| Rurality | 1.24 | 0.179 | 0.128 | 0.939 | 1.650 |
| Job Disruption | 1.09 | 0.157 | 0.548 | 0.823 | 1.444 |
| Survey Year | 1.09 | 0.154 | 0.553 | 0.824 | 1.437 |

Table S30. Ordinal logistic regression results predicting the effects of hunting on frequency of white meat intake, by food security status. Statistically significant ($p<0.05$) results are bolded for emphasis.

| **White Meat Intake Frequency** | **Odds Ratio** | **Standard Error** | **p-value** | **95% CI** | |
|---|---|---|---|---|---|
| | | | Food Secure | | |
| Hunting | 1.25 | 0.183 | 0.134 | 0.935 | 1.660 |
| Race/Ethnicity | 0.82 | 0.210 | 0.448 | 0.499 | 1.359 |
| Gender Identity | 1.15 | 0.134 | 0.238 | 0.913 | 1.442 |
| **Age** | **0.73** | **0.085** | **0.006** | **0.577** | **0.913** |
| Education Level | 0.85 | 0.101 | 0.168 | 0.673 | 1.071 |
| **Household Income** | **1.56** | **0.193** | **0.000** | **1.226** | **1.989** |
| Rurality | 1.02 | 0.114 | 0.858 | 0.819 | 1.271 |
| **Job Disruption** | **1.49** | **0.196** | **0.003** | **1.147** | **1.925** |
| Survey Year | 0.87 | 0.097 | 0.223 | 0.704 | 1.085 |
| | | | Food Insecure | | |
| **Hunting** | **1.75** | **0.354** | **0.006** | **1.177** | **2.602** |
| Race/Ethnicity | 0.67 | 0.181 | 0.135 | 0.392 | 1.135 |
| Gender Identity | 0.87 | 0.131 | 0.355 | 0.647 | 1.169 |
| **Age** | **0.61** | **0.150** | **0.043** | **0.373** | **0.983** |
| Education Level | 1.06 | 0.163 | 0.709 | 0.783 | 1.433 |
| **Household Income** | **1.44** | **0.238** | **0.026** | **1.045** | **1.994** |
| **Rurality** | **1.41** | **0.206** | **0.020** | **1.054** | **1.873** |
| Job Disruption | 1.13 | 0.158 | 0.399 | 0.855 | 1.483 |
| Survey Year | 1.20 | 0.174 | 0.217 | 0.900 | 1.591 |
